# Your Heart Failure Prediction to Identify Un-diagnosed Patients from Routine Primary Care Records

**DOI:** 10.1101/2025.10.02.25337031

**Authors:** Cindy Leyvraz, Zeina Gabr, Eva Sarlin, Nouchine Hadjikhani, André Jaun

## Abstract

**Background and Aims:** Heart Failure is a common and serious condition that often remains undetected until a major cardio-vascular event leads to diagnosis is secondary care. Here we propose a portable artificial intelligence tool that integrates clinical guidelines with phenotypic markers to identify high-risk patients who may benefit from formal diagnosis evaluation and timely initiation of treatment.

**Methods:** Diagnosis guidelines are first encoded using a rule-based model, which is then used to train a neural network. Relying on de-identified real-world evidence from UK primary care, transfer learning is used to train on 91,346 historical records and forecast the 6.2% patients who received a diagnosis within 3 years. Tested for portability in an independent sample consisting of 56,308 validation records, predictions are interpreted using Shapley values and individually assessed for statistical significance by comparison with matched digital twin cohorts. A Kaplan-Meier survival analysis links positive predictions to the observed excess mortality.

**Results:** Compared with the prevailing challenge of under-diagnosis, model predictions in the validation set (0.7% TP, 2.7% FP) demonstrate strong statistical support, with fewer than 1.5% failing to reject a null hypothesis at p=0.05. Among the TP, the likelihood of receiving a future diagnosis is over 7.6 times higher than the baseline prevalence in the validation cohort. In both TP and FP cohorts, patients aged 60-70 years exhibited mortality rates more than fivefold higher than the control population. Furthermore, variables derived from the Complete Blood Count (CBC) including white blood cell count (WBC) and red cell distribution width (RDW), contribute significant predictive value beyond established diagnosis criteria.

**Conclusions:** When implemented within a clinical decision support system, predictive AI has the potential to improve patients outcomes by leveraging routinely collected phenotypic markers, which are challenging for clinicians to interpret in the context of complex decision-making pathways.^1^

## Introduction

Heart failure (HF) is a widespread, life-threatening condition that frequently goes undiagnosed until patients present with severe symptoms or suffer a major cardiovascular event requiring hospital care. In England, as many as 80% of HF cases are only first identified in hospitals [Butler08, Bottle18]. Early identification in primary care is hindered by the disease’s often subtle and nonspecific clinical presentation such as fatigue, and the complexity of diagnostic criteria [Nadarajah24, Medscape25]. It may even be more challenging to detect in older individuals with other overlapping conditions. Conventional approaches therefore risk missing patients in the early or preclinical stages.

To address this challenge, we introduce a portable artificial intelligence (AI) tool designed to support clinical decision-making by identifying individuals at high risk of HF. The system integrates both established diagnostic guidelines and phenotypic markers extracted from routinely collected health records [Foy25].

A neural network is first trained on clinical guidelines and then refined using real-world evidence from UK primary care, enabling it to forecast patients who may require immediate diagnosis or who are likely to develop HF within three years. Importantly, the model exploits predictive signals from routine investigations such as Complete Blood Count (CBC), which are often underutilized in clinical practice, yet carry significant forecasting power.

## Methods

To develop a portable and data efficient tool, clinical guidelines are first encoded in a decision tree and used to train a neural network that accurately replicates [NICE NG106] heart failure diagnosis criteria. Leveraging anonymised data from two widely adopted and distinct electronic medical record systems [EmisHealth25, SystmOne25], the model is initially retrained on real-world evidence (RWE) from the first dataset to predict diagnoses up to 3 years in advance. The second dataset is reserved for validation, where model forecasts are evaluated using Shapley values [Shapley51]. This approach enables rigorous comparison with digital twin cohorts, generating statistically significant factual and counterfactual evidence in the retrospective analysis.

Diagnostic guidelines can be represented as a rule-based model relying on coded symptoms (such as Shortness of breath, Orthopnea, Paroxysmal nocturnal dyspnoea, Reduced exercise tolerance, Tiredness, Fatigue, Dizziness, Ankle oedema, Ankle swelling, Palpitations, Weakness, Wheezing, Abdominal distension, Leg swelling, Nocturnal cough, Weight gain, Weight loss, Fluid retention, Pulmonary oedema, Irregular pulse, Tachycardia, Odema, Resting sinus tachycardia, Elevated jugular venous pressure, Third heart sound, Cardiac murmur, Ascites, Pleural effusion, Peripheral oedema, Scrotal oedema, Hepatomegaly, Supraventricular tachycardia, Cardiomegaly, Abnormal echocardiogram, Abnormal chest X-ray and Abnormal ECG), lab results (NT-ProBNP) to determine a formal diagnosis or refer to specialist.

A feed-forward neural network^2^ with a pyramidal layout of (25, 17, 10) neurons is initially trained with synthetic data to reproduce diagnosis rules with more than 95.7% accuracy. Missing features are set to 0, with a sentinel feature added to account for their absence. Additional factors with forecasting power are initialized with white noise:

- age
- Body Mass Index (BMI)
- Systolic Blood Pressure (SBP)
- Estimated Glomerular Filtration Rate (eGFR)
- Glycated Hemoglobin (A1c)
- Alanine Aminotransferase (ALAT)
- Aspartate Aminotransferase (ASAT)
- Urinary Albumin-to-Creatinine Ratio (Alb/Creat)
- B-type Natriuretic Peptide (BNP)
- C-Reactive Protein (CRP)
- Creatinine
- Heart Rate (HR)
- Low-Density Lipoprotein Cholesterol (LDLc)
- Blood Sodium Level (Na)
- Red Cell Distribution Width (RDW)
- Mean Corpuscular Hemoglobin Concentration (MCHC)
- Random Plasma Glucose (RPG)
- Serum Albumin (Serum_Alb)
- Blood Urea (Urea)
- White Blood Cell Count (WBC)
- Gender

One of the key challenges in learning from RWE data is the prevalence of missing diagnoses – many patients live with the condition without ever receiving a formal diagnosis [Bottle18, Hancock12]. This results in biased training data that underrepresent true cases. To mitigate this issue and reduce the impact of diagnosis underreporting, we implement a corrective strategy aimed at improving the robustness and generalizability of the training data by discarding negative evidence for records having at least one cardio-metabolic diagnosis.

The training data set stems from 91,346 records, of which 5,564 (6.2%) have a formal HF diagnosis code. Using multiple evaluations for every history and a 3 years look-ahead to detect subsequent diagnosis, this results in 747,524 training samples, of which 22,598 (3.0%) are labelled as developing a HF condition.

For each prediction, explanations are provided through Shapley values [Shapley51; Lundberg17] and through its “digital twins”, defined as clinically similar cases corresponding to neighboring predictions in Shapley space measured by Manhattan distance across all factors. To evaluate whether predictions are grounded in clinical evidence rather than model ‘hallucination,’ both statistical support and counterevidence are assessed by comparing the proportion of digital twins diagnosed with heart failure within 3 years against the baseline proportion in the validation set, using a one-tailed z-test at *p*=0.05.

Usability testing was carried out by a medical doctor who accessed 84 records from a pool of 415 undiagnosed patients aged 65+ years, by reviewing how useful a HF forecast is in the light of the existing conditions. The recommendations for individual patients were displayed in a clinical decision support tool with Shapley value explanations and p-value testing with an example shown in figure 1.

**Figure 1.**
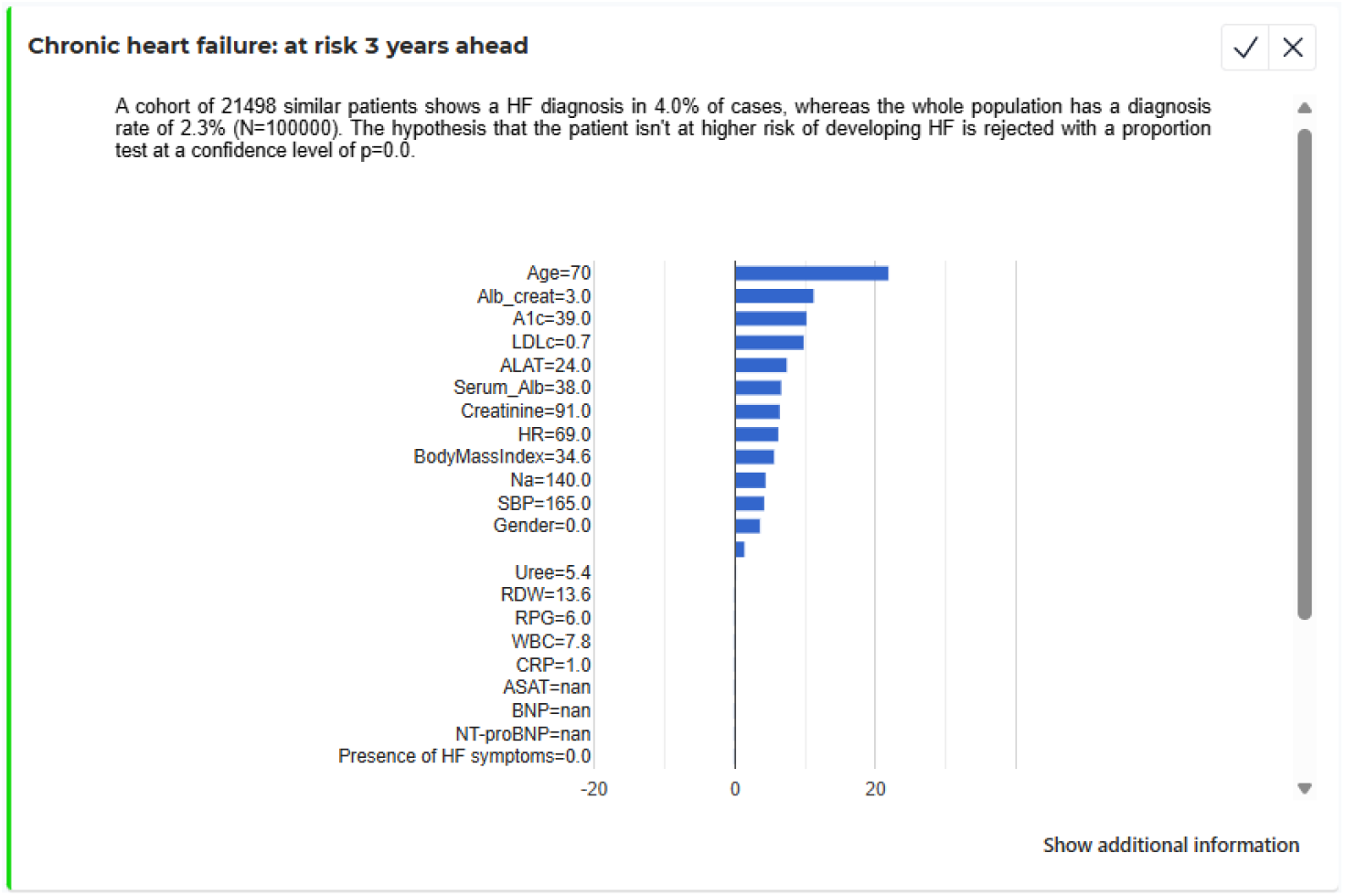
Display from the usability testing supporting clinicians to identify heart failure risk for one specific patient, explaining the forecast with Shapley values and rejecting the hypothesis test with p<0.005.

**Figure 2.**
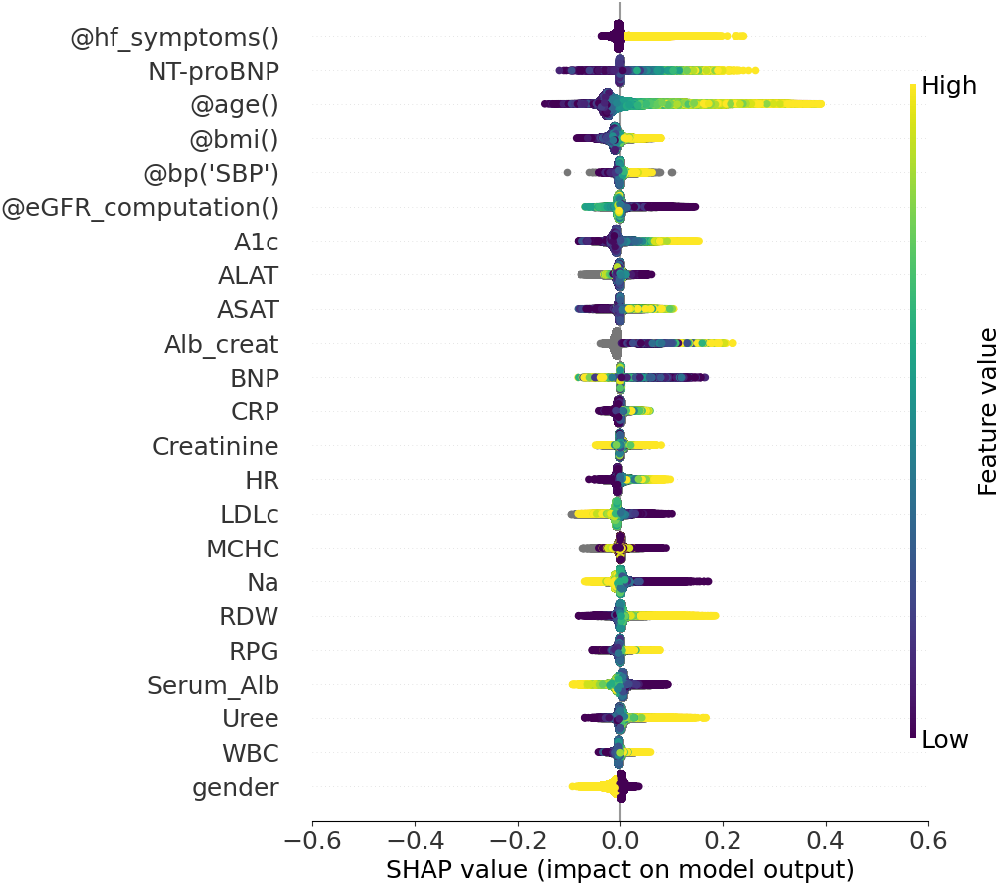
SHAP summary plot for the predictions in the validation dataset, displaying the overall feature importance (x-axis) and correlation with feature value (color).

The adequacy of the recommendation was evaluated in the light of the broader context of medical conditions and categorized from 1-3 accept (insightful, unsurprising, trial to consider) to 4-6 reject (too risky, inadequate, dangerous).

## Results

For a fair assessment of the forecasting capability across different clinical settings, the validation data stem from a separate electronic medical records system that has not been used for training. A total of 56,308 patient historical records, of which 3,060 (5.4%) have a HF diagnosis, are used to create 486,707 samples where AI-driven recommendations are assessed against a diagnosis appearing within 3 years from the forecast. About one fifth of the diagnoses are recognized early (0.7% true positives, TP) and five times more overlap with a potentially underdiagnosed population (2.7% false positive, FP) where “false” means that patients were never diagnosed, but may have been overlooked by human clinicians. The complete confusion matrix reads 3,399 (0.7%) TP, 13,020 (2.7%) FP, 8,054 (1.6%) FN and 462,234 (95.0%) TN.

Figure 1 illustrates how different features contribute to the explanation: large values of age, NT-proBNP and multiple HF symptoms are unsurprisingly linked with an increased probability of HF forecasts, but a much wider range of features (notably RDW, Urea, A1c, ASAT and RPG) contribute and often dominate the prediction in aggregate, suggesting that it is difficult to replace the neural network with cohort specific analytical models.

Deep neural networks are well known to “hallucinate” when the inference draws from sparsely populated regions of the parameter space.To guard against this, forecasts are validated using statistical evidence independent of AI. Figure 3 illustrates this validation by reporting the proportion of diagnoses confirmed within 3 years, providing both factual (similar “digital twin” patients with similar outcomes) and counterfactual (dissimilar patients with different outcomes) evidence. These results are shown for “digital twin” cohorts constructed around confirmed diagnoses (0.7% TP), using hyperspheres in Shapley value space centered on each forecast, with progressively increasing radii. As the radius expands, the cohort enlarges but becomes progressively less representative of the original prediction, ultimately encompassing the entire population.

**Figure 3.**
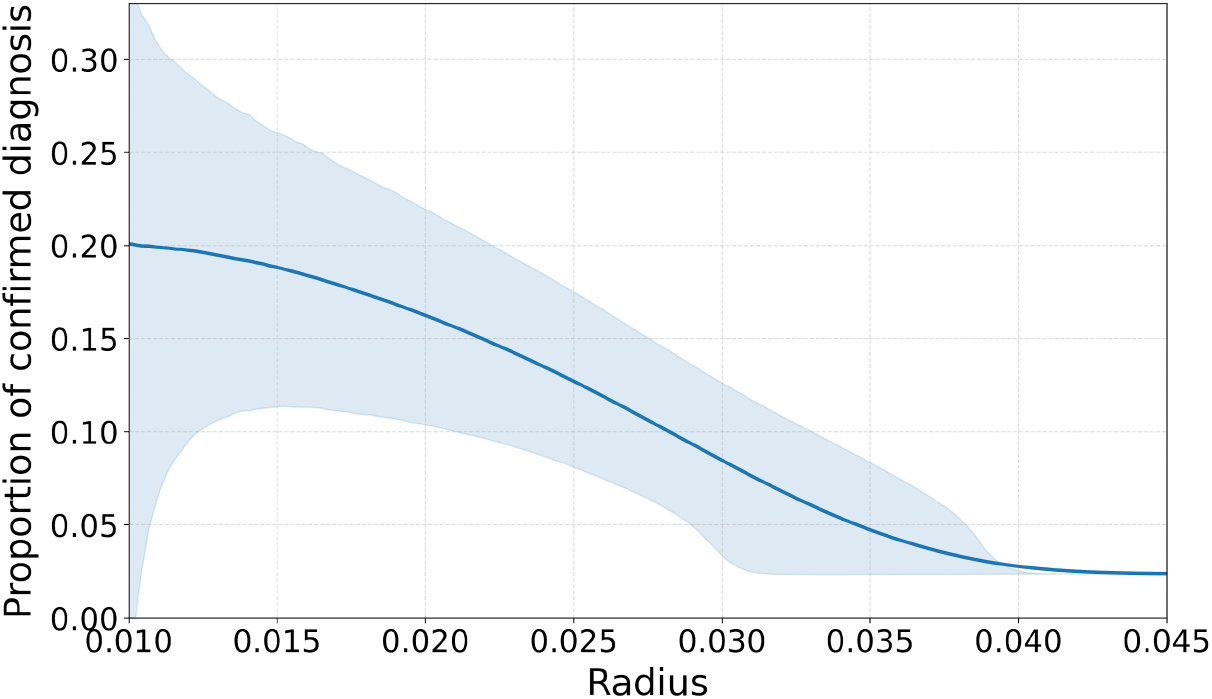
Proportion of HF forecasts confirmed by a clinician within 3 years as a function of case similarity. As neighbors become less similar in Shapley value space, the mean and 68% percentile bands decline, converging toward the validation set baseline (0.023). This pattern provides both factual and counterfactual statistical evidence of forecasting capability.

Small radii correspond to narrowly defined digital twin cohorts, which exhibit a wider variability in outcomes due to statistical noise, but with an average of ∼20% - reflecting the fraction of clinically similar patients who received a confirmed diagnosis within 3 years. At larger radii, the proportion converges towards the overall prevalence of HF diagnosis in the validation set (2.3%). Using clinician-confirmed diagnoses as a reference standard, Figure 3 demonstrates that the AI isolates cohorts characteristics associated with a tenfold higher likelihood of diagnosis compared to the general population, and that in the absence of these characteristics, the likelihood reverts to the population baseline.

Because a substantial number of cases may remain undiagnosed in the clinic, we further characterize the unconfirmed cohort (2.7% FP) and examine its relationship to disease progression. In the absence of a universally available biomarker, but leveraging mortality as a severity endpoint, we conducted a Kaplan-Meier survival analysis comparing mortality rates with those of the general patient population. As shown in Table 1, mortality is elevated in both confirmed (TP) and unconfirmed (FP) cohorts, with the excess risk particularly pronounced - approximately fivefold - in younger patients aged 60-65 years. Although the effect attenuates with increasing age, it remains more than twofold higher in patients up to 75 years old.

**Table 1.** Kaplan-Meier analysis of the 10 years mortality observed for HF forecasts in comparison with the entire population (Control), suggesting that both TP (0.7% who are diagnosed within 3 years) and FP (2.7% who are not) have similar survival rates and significant excess mortality in particular for the younger age groups.

| Age [yr] | Cohort | N | Deaths | 10y Survival | Mortality [%/yr] | Excess Mortality |
| --- | --- | --- | --- | --- | --- | --- |
| 60–65 | Control | 13,188 | 261 | 0.98 | 0.4 | – |
|  | FP | 210 | 22 | 0.895 | 2.1 | 5.3× |
|  | FP + TP | 232 | 22 | 0.905 | 1.9 | 4.8× |
| 65–70 | Control | 11,423 | 438 | 0.962 | 0.77 | – |
|  | FP | 584 | 76 | 0.826 | 2.6 | 3.4× |
|  | FP + TP | 647 | 86 | 0.867 | 2.66 | 3.5× |
| 70–75 | Control | 9,073 | 592 | 0.935 | 1.3 | – |
|  | FP | 1,196 | 163 | 0.864 | 2.73 | 2.1× |
|  | FP + TP | 1,343 | 189 | 0.859 | 2.81 | 2.2× |
| 75–80 | Control | 5,886 | 741 | 0.874 | 2.52 | – |
|  | FP | 1,254 | 257 | 0.795 | 4.1 | 1.6× |
|  | FP + TP | 1,472 | 280 | 0.81 | 3.8 | 1.5× |

A retrospective analysis cannot formally establish a causal relationship between the likely onset of HF and the observed mortality. Nevertheless, the combination of factual and counter-factual evidence (Fig. 3, Table 1), together with the forecast of an outcome exogenous to the model (mortality), provides sufficient justification for collecting additional evidence (NT-proBNT, LVEF) to support a potential diagnosis.

From a regulatory perspective, clinical deployment is further supported by usability testing, which showed that none of the AI-generated recommendations were deemed too risky, inadequate, or dangerous, one was accepted with caution, and 64 accepted without reservation.

## Conclusion

Our study highlights the transformative potential of an AI model to enable earlier detection of heart failure, improve patient outcome, and strengthen clinical decision-making, even in complex care settings with limited data availability.

## Data Availability

All data produced in the present work are contained in the manuscript.

## Footnotes

1 One liner. A neural network is trained on clinical guidelines and fine-tuned on UK primary care data to identify patients at high risk of requiring an immediate diagnosis of developing heart failure condition within three years.

2 nnid: 6854265245792384d3654549 (guidelines), 6892147f0c1abc50682e301b (tl)

